# Blood *N*-glycomic signature of fibrosis in MASLD shows low levels of global α2,3-sialylation

**DOI:** 10.1101/2024.09.19.24313949

**Authors:** Tamas Pongracz, Bart Verwer, Anne Linde Mak, Oleg A. Mayboroda, Simone Nicolardi, Marco R. Bladergroen, Wenjun Wang, Maaike Biewenga, Max Nieuwdorp, Joanne Verheij, Adriaan G. (Onno) Holleboom, Bart van Hoek, Noortje de Haan, Manfred Wuhrer, Maarten E. Tushuizen

**Author notes:** Shared senior and corresponding authors: Manfred Wuhrer; Maarten Tushuizen.

## Abstract

**Background & Aims:** Alterations in the glycosylation of blood proteins affect protein functionality and have been linked to various diseases. Metabolic dysfunction-associated steatotic liver disease (MASLD) is a silent disease, of which progression to advanced disease stages including metabolic dysfunction-associated steatohepatitis (MASH), fibrosis and cirrhosis often goes unnoticed. As current non-invasive diagnostic tests lack specificity, the purpose of this work was to study total blood protein *N*-glycosylation in individuals with MASLD and various degrees of fibrosis as compared to healthy controls.

**Methods:** In two independent cross-sectional cohort studies, blood *N*-glycosylation analysis was performed by mass spectrometry on released glycans of overall 132 MASLD patients and 99 age- and sex-matched healthy controls. Relationships between glycosylation traits and the disease spectrum of MASLD including fibrotic MASLD were investigated in comparison to healthy controls. Furthermore, publicly available transcriptomics datasets were used to explore glycosyltransferase expression in patients with MASLD.

**Results:** Globally lower α2,3-sialylation distinguished MASLD from healthy controls (OR [CI]=0.36; [0.18-0.67]; *p*-value=0.019, and 0.11 [0.04-0.24]; *p*-value<0.000001), as well as non-fibrotic MASLD from its fibrotic counterparts (OR: 0.13 [0.06-0.26]; *p*- value<0.0001), but showed no association with steatohepatitis activity. Hepatic *ST3GAL6*, a sialyltransferase responsible for *N*-glycan α2,3-sialylation, negatively associated with fibrosis progression, similar to the observed glycomic signature. Both glycomic and transcriptomic signatures were replicated in independent cohorts.

**Conclusions:** Fibrotic MASLD is characterized by a global decrease of blood protein α2,3-sialylation and according decrease in hepatic α2,3-sialyltransferase expression, associating with disease progression. These findings suggest alterations in the *N*- glycan biosynthetic pathway and are potentially useful in the early diagnosis of fibrosis in MASLD.

**Lay Summary:** Non-invasive markers of fibrotic MASLD perform suboptimal. This research identified that changes in blood protein glycosylation coincide with fibrosis development, offering blood-based markers that could potentially replace a liver biopsy.

**What You Need to Know:** *BACKGROUND AND CONTEXT:* The majority of the plasma glycoproteins is synthesized in the liver and changes to their glycosylation are known to affect their function and to associate with liver disorders.

*NEW FINDINGS:* MASLD patients exhibit lower α2,3-sialylation on the complete range of their blood protein N-glycans, which coincides with the histological appearance of fibrosis, mediated likely via downregulation of hepatic *ST3GAL6*.

*LIMITATIONS:* While the findings of this study has could have implications for diagnosing fibrotic MASLD, the identified glycomic signature needs to be confirmed in a larger, ideally prospective patient cohort.

*CLINICAL RESEARCH RELEVANCE:* By identifying specific signatures in the blood protein N-glycome, this research offers potential non-invasive markers for early diagnosis and monitoring of fibrosis in MASLD. Non-invasive diagnosis could potentially lessen the need for liver biopsies, and allow for timely intervention and improved disease management, ultimately leading to improvement of patient outcomes and the reduction of liver-related morbidity and mortality.

*BASIC RESEARCH RELEVANCE:* The observed glycomic and transcriptomic signatures offer molecular-level insights into fibrosis development in MASLD. This paves the way for future research at the intersection of glycoscience and hepatology, that will offer deeper insights into the pathophysiology of this liver disease.

## Introduction

Metabolic dysfunction-associated steatotic liver disease (MASLD; previously known as non-alcoholic fatty liver disease (NAFLD)) is a disease of alarmingly increasing prevalence, linked to metabolic, cardiovascular and malignant morbidity^1, 2^. It is a spectrum of liver disease, ranging from isolated steatosis, in which the predominant histological characteristic is lipid accumulation in hepatocytes, to metabolic-dysfunction associated steatohepatitis (MASH; previously known as non-alcoholic steatohepatitis (NASH)). MASH is characterized by additional hepatic inflammation, in turn triggering a maladaptive repair response, which causes incremental stages of fibrosis, ultimately culminating in MASH-related cirrhosis and hepatocellular carcinoma (HCC)^1^. Obesity and insulin resistance are strongly associated with MASLD, both via increased delivery of free fatty acids to the liver and through increases of hepatic lipogenesis associated with hyperglycemia and hyperinsulinemia^2^. With the global increase in obesity and insulin resistance/type 2 diabetes mellitus (T2DM), MASLD has become the most prevalent liver disease in the world, occurring in up to 25-30% of adult populations^3, 4^.

Increased liver fibrosis has been identified as the only MASLD factor associated with higher overall and liver-related mortality, as well as a greater likelihood of developing liver-related complications, including HCC and the need for liver transplantation. In contrast, steatosis grade or inflammatory disease activity itself does not correlate with these outcomes^5, 6^. This underscores the importance of fibrosis staging in the clinical assessment of the severity of MASLD, to ensure initiation of appropriate and timely multidisciplinary treatment plans. These treatment plans may include lifestyle and dietary interventions, participation in clinical trials, consideration of bariatric surgery and since very recently pharmacotherapy in the form of resmetirom^1, 7, 8^.

A liver biopsy remains the clinical reference standard for detection and staging of MASLD fibrosis, but has the potential for complications and suffers from sampling and reading errors^9, 10^. Radiologic assessment of liver stiffness has gained credibility over the past decade as a diagnostic and staging tool for liver fibrosis with vibration-controlled transient elastography (i.e., FibroScan) and magnetic resonance imaging^11,12^. Simple composite proxies of liver fibrosis, including FIB-4 (age, platelet count, aminotransferases), NAFLD fibrosis score (NFS), and serologic panels of combined biomarkers like Enhanced Liver Fibrosis (ELF), have good accuracy in excluding advanced fibrosis, and could be used to identify individuals at low risk for advanced disease in high-risk populations^12–14^. However, their implementation in primary care is not well established. Their relatively high rates of false negative and false positive outcomes in general populations may make their use suboptimal^15, 16^.

Glycans are a class of biomarkers that have been reported to convey promising diagnostic and prognostic potential in the context of liver pathologies^17, 18^. Most blood proteins are synthesized and glycosylated in the liver, and modifications to the glycan moiety are known to affect their structure and function^19^. Such alterations have been linked to liver diseases and conditions such as MASLD-MASH and corresponding fibrosis stages^20–27^, cirrhosis^23, 28^, HCC^29, 30^, alcoholic liver disease^31^, and more recently autoimmune hepatitis (AIH)^32^. In turn, particular congenital forms of severe steatohepatitis resulting from defective turnover of hepatocyte lipid droplets also present with defective plasma protein glycosylation patterns^33, 34^.

Protein glycosylation is a non-template-based, tightly regulated biosynthetic process, the regulation of which depends on environmental, metabolic, (epi)genetic and cellular factors^19, 35^. The latter include the availability of glycosyltransferases and substrates^36^. Glycosylation patterns can serve as distinct signatures that reflect cellular differentiation phenotypes as well as various physiological and pathological events^37–39^, and have been found to be paralleled by a directionally similar change in the expression of glycosyltransferases involved in their biosynthesis^40–43^.

Here, we employed a semi-automated high-throughput mass spectrometry-based glycomics approach to explore blood protein *N*-glycosylation on the released glycan level, i.e., after enzymatic liberation of *N*-glycans from their carrier proteins. We identified a biomarker allowing for early, non-invasive detection of fibrotic MASLD. This marker has the potential to meet the clinical need of capturing non-fibrotic to fibrotic transition immediately when histological signs appear, and is promising as a tool in primary care as glycans are amenable for dried blood spot-based sample collection and analysis^44^.

## Materials and methods

All authors had access to the study data and reviewed and approved the final manuscript.

### Study design

Samples were obtained from the biobanks of Leiden University Medical Center (LUMC; discovery and replication cohort) and Amsterdam University Medical Center (replication cohort part, ANCHOR^12^). The discovery cohort (LUMC) involved 30 patients with MASLD (non-invasively diagnosed by imaging including ultrasound and MRI) and 60 presumably healthy, age- and sex-matched controls with plasma samples. The combined replication cohorts (LUMC+ANCHOR) included 102 patients with MASLD and 29 presumably healthy, age- and sex-matched controls with serum samples. Samples were included based on availability. The cohort descriptions can be found in **Table 1**. The study protocol was approved *a priori* by the local ethical committees (B19.071 and B21.045) for the discovery and replication cohort, respectively. Healthy controls in the replication cohort were obtained via the LUMC voluntary donor service (“LUMC Vrijwillige Donoren Service”). Informed consent was obtained from all patients and healthy controls, and the studies complied with the latest version of the Declaration of Helsinki.

**Table 1.** Demographic and clinical characteristics of patients and healthy controls in the discovery and replication cohorts.

| Cohort | Discovery |  | Replication |  |  |  |  |  |  |
| --- | --- | --- | --- | --- | --- | --- | --- | --- | --- |
| Group (n) | Healthy (60) | MASLD (30) | Healthy (29) | MASLD (102) | Fibrosis stage (Brunt) |  |  |  |  |
|  |  |  |  |  | F0 (12) | F1 (15) | F2 (37) | F3 (14) | F4 (24) |
| Sex (male n, %) | 17 (28%) | 13 (43%) | 19 (66%) | 64 (63%) | 6 (50%) | 7 (46.7%) | 27 (73%) | 7 (50%) | 17 (70.8%) |
| Age (mean, SD) | 49 (17) | 52 (17) | 45.3 (13.4) | 51.1 (14.7) | 47 (17.2) | 48 (15) | 48 (15) | 56 (14) | 58 (11) |
| BMI (mean, SD) |  |  |  | 31.3 (7.6) | 28.2 (10.6) | 32.2 (4.0) | 33.1 (5.3) | 32.2 (4.6) | 29.3 (10.7) |
| T2DM (n, %) |  |  |  | 49 (48%) | 1 (8.3%) | 6 (40%) | 14 (37.8%) | 9 (64.3%) | 19 (79.2%) |
| Hypertension (n, %)* |  |  |  | 22 (21.6%) | 0 (0%) | 4 (26.7%) | 4 (10.8%) | 2 (14.3%) | 12 (50%) |
| AST (mean, SD) |  | 33 (18.2) |  | 59.8 (54.3) | 34.6 (12.0) | 54.7 (33.1) | 58.3 (38.7) | 63.1 (29.6) | 76.0 (93.2) |
| ALT (mean, SD) |  | 55.9 (40.7) |  | 70.0 (60.6) | 51.4 (26.1) | 101.3 (83.2) | 75.7 (48.7) | 57.5 (43.9) | 58.4 (74.9) |
| Triglycerides* (mean, SD) |  |  |  | 1.5 (1.9) | 1.2 (0.9) | 1.8 (1.3) | 1.6 (1.4) | 1.4 (0.9) | 1.5 (2.6) |
| FIB4 (mean, SD) |  |  |  | 2.7 (3.9) | 0.9 (0.5) | 1.2 (0.8) | 1.4 (1.2) | 2.0 (0.9) | 6.9 (6.4) |
| NFS (mean, SD) |  |  |  | -0.8 (2.3) | -3.1 (2.6) | -1.5 (1.9) | -1.8 (1.4) | 0.0 (1.4) | 1.8 (1.5) |
| NAS $\geq 4$ (n, %) | | | | 59 (57.8%) | 4 (33.3%) | 7 (46.7%) | 26 (70.3%) | 11 (78.6%) | 11 (45.8%) |
| Steatosis (n, % by row) |  |  |  | 90 |  |  |  |  |  |
| 0 |  |  |  | 7 | 2 (28.6%) | 0 (0%) | 3 (42.9%) | 0 (0%) | 2 (28.6%) |
| 1 |  |  |  | 36 | 5 (13.9%) | 8 (22.2%) | 8 (22.2%) | 4 (11.1%) | 11 (30.6%) |
| 2 |  |  |  | 26 | 2 (7.7%) | 2 (7.7%) | 17 (65.4%) | 3 (11.5%) | 2 (7.7%) |
| 3 |  |  |  | 21 | 0 (0%) | 5 (23.8%) | 8 (38.1%) | 6 (28.6%) | 2 (9.5%) |
| **Lobular inflammation (n, % by row)** |  |  |  | 89 |  |  |  |  |  |
| 0 |  |  |  | 7 | 2 (28.6%) | 3 (42.9%) | 0 (0%) | 0 (0%) | 2 (28.6%) |
| 1 |  |  |  | 71 | 4 (5.6%) | 10 (14.1%) | 32 (45.1%) | 12 (16.9%) | 13 (18.3%) |
| 2 |  |  |  | 11 | 3 (27.3%) | 2 (18.2%) | 4 (36.4%) | 1 (9.1%) | 1 (9.1%) |
| **Portal inflammation (n, % by row)** |  |  |  | 84 |  |  |  |  |  |
| 0 |  |  |  | 28 | 5 (17.9%) | 6 (21.4%) | 10 (35.7%) | 3 (10.7%) | 4 (14.3%) |
| 1 |  |  |  | 35 | 3 (8.6%) | 6 (17.1%) | 18 (51.4%) | 4 (11.4%) | 4 (11.4%) |
| 2 |  |  |  | 20 | 1 (5%) | 2 (10%) | 5 (25%) | 5 (25%) | 7 (35%) |
| 3 |  |  |  | 1 | 0 (0%) | 0 (0%) | 0 (0%) | 0 (0%) | 1 (100%) |
| **Ballooning (NAS; n, % by row)** |  |  |  | 90 |  |  |  |  |  |
| 0 |  |  |  | 30 | 6 (20%) | 7 (23.3%) | 12 (40%) | 1 (3.3%) | 4 (13.3%) |
| 1 |  |  |  | 40 | 2 (5%) | 7 (17.5%) | 20 (50%) | 6 (15%) | 5 (12.5%) |
| 2 |  |  |  | 20 | 1 (5%) | 1 (5%) | 4 (20%) | 6 (30%) | 8 (40%) |

\*Incomplete observations

### Liver biopsy and assessment of fibrosis

In the replication cohort, fibrosis scores were defined according to the Brunt scoring system^45^, based on histological examination of liver biopsies by two independent pathologists as follows: F0: no fibrosis (no scarring); F1: portal fibrosis (minimal scarring); F2: periportal fibrosis (significant scarring has occurred and extends outside the liver area); F3: severe fibrosis (fibrosis spreading and forming bridges with other fibrotic liver areas); F4: cirrhosis (advanced scarring). No such fibrosis-specific readout was available for patients enrolled in the discovery cohort, although the presence of cirrhosis, affecting 4 patients, was defined on the basis of histology or if unavailable, using liver elastography or liver ultrasound. Decompensated liver cirrhosis was defined as presence of ascites, varices bleeding, HCC, hepatorenal or hepatopulmonary syndrome.

### Mass spectrometry glycomics and data processing

The total blood *N*-glycome was analyzed by matrix-assisted laser desorption/ionization – Fourier-transform ion cyclotron resonance – mass spectrometry (MALDI-FTICR-MS) after linkage-specific sialic acid derivatization, as described previously^46^ **(Supplementary Materials & Methods)**. After initial data pre- processing including data quality control, similarly to preceding reports^47^ **(Supplementary Materials & Methods)**, the relative abundances of individual glycans were calculated by normalizing them to their total area **(Supplementary Table 1, 2)**. Glycosylation traits summarizing specific glycosylation features that reflect biosynthetic pathways were calculated using the relative abundance of individual glycans **(Figure 1, Supplementary Table 3, 4)**.

**Figure 1.**
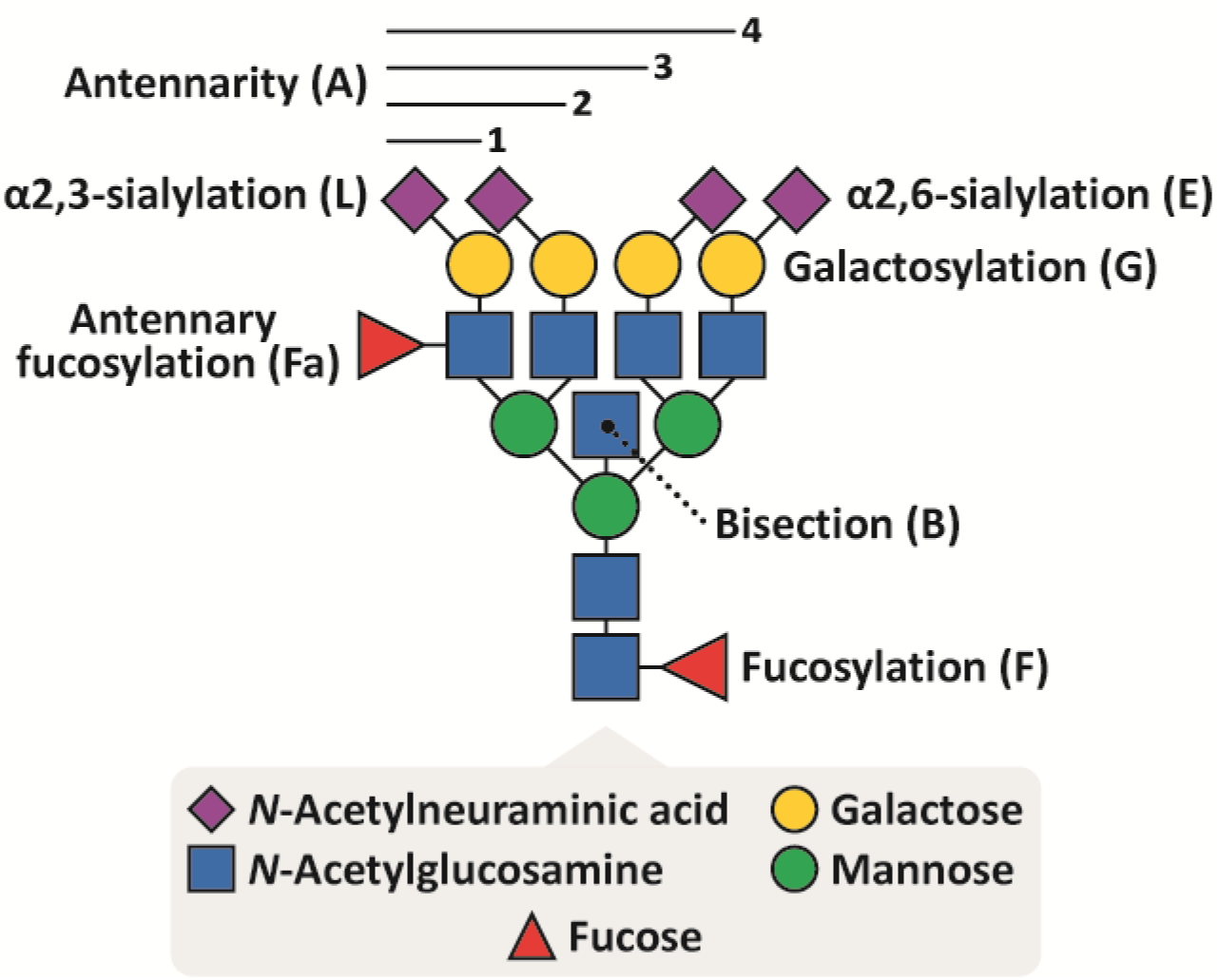
Monosaccharide constituents that build up human *N*-glycans (bottom) and glycosylation traits calculated therefrom, as illustrated on a fully sialylated tetraantennary *N*- glycan (top).

### Re-analysis of publicly available RNA-seq data

Two publicly available total RNA-seq datasets on biopsy-obtained liver tissue sections were used to assess sialyltransferase transcript expression patterns across the fibrotic MASLD spectrum (Gene Expression Omnibus accession numbers GSE162694 (Cohort 1) and GSE130970 (Cohort 2))^48, 49^. For cohort characteristics, please see **Supplementary Table 5** and the cited works. Prior to analysis, counts were scaled by the total number of reads for each gene (counts per million normalization), followed by log-transformation of the obtained normalized values.

### Statistical analysis

For both glycomics cohorts, a logistic regression model on standardized data (subtraction of the mean and division by the SD) including age, sex and their interaction as co-variates was used to study the associations between glycosylation of healthy controls and patients with MASLD (Healthy=0; MASLD=1) **(Table 2, Supplementary Table 6, Supplementary Materials & Methods)**. Additionally, a principle component analysis (PCA) was performed to reveal potential drivers of separation between healthy and diseased, as well as any potential batch effects, showing that the separation of standards was not driven by systematic batch effects **(Supplementary** Figure 1)^32^. To compare how the significantly different glycosylation traits, or glycosyltransferase expression levels, differ between groups with varying Brunt fibrosis score, a Kruskal-Wallis test was performed, which in case of a significant result, was followed by the post-hoc Dunn’s test **(Supplementary Table 7-10; Figure 3**, **Figure 5)**. Spearman’s ranked correlation was performed to assess the correlation of conventional diagnostic markers, glycosylation traits and glycosyltransferase expression levels with Brunt fibrosis score as well as with MASLD activity (NAFLD activity score (NAS score), steatohepatitis, lobular and portal inflammation, ballooning), age and BMI **(Table 3, Supplementary** Figure 7**, Supplementary Table 11)**. To account for multiple testing, during the evaluation of statistical significance per statistical question, the Benjamini-Hochberg procedure with a false discovery rate of 5% was used **(Figure 2** and **Table 2** (discovery cohort), **Figure 4**, **Supplementary Table 6, 8, 10, 11**). However, statistical testing in the replication cohort and in correlation analyses was performed without multiple testing correction using a cut-off of *p* < 0.05 **(Table 2-3, Supplementary Table 6, Supplementary** Figure 7).

**Figure 2.**
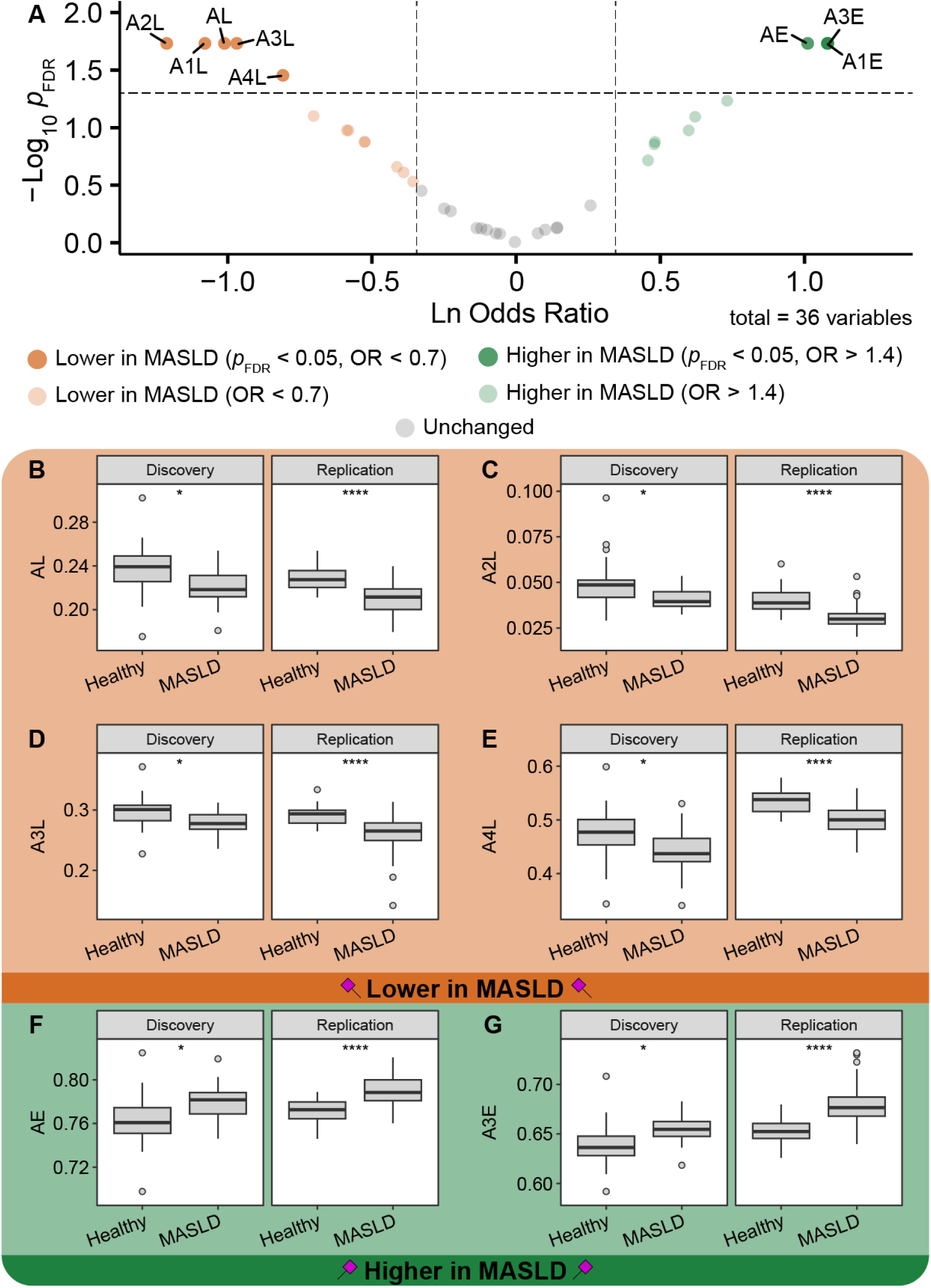
Replicated associations between patients with MASLD and healthy controls. **(a)** Volcano plot based on the calculated 36 glycosylation traits in the discovery cohort. **(b-g)** Comparison of the relative abundance differences between healthy controls and MASLD showing the replicated glycosylation traits with negative **(b, e)** and positive **(f, g)** effect sizes. *P*-values, ORs and 95% CIs are shown in **Table 2** and **Supplementary Table 6**. *, **: *p*-value < 0.05, 0.01, respectively.

**Figure 3.**
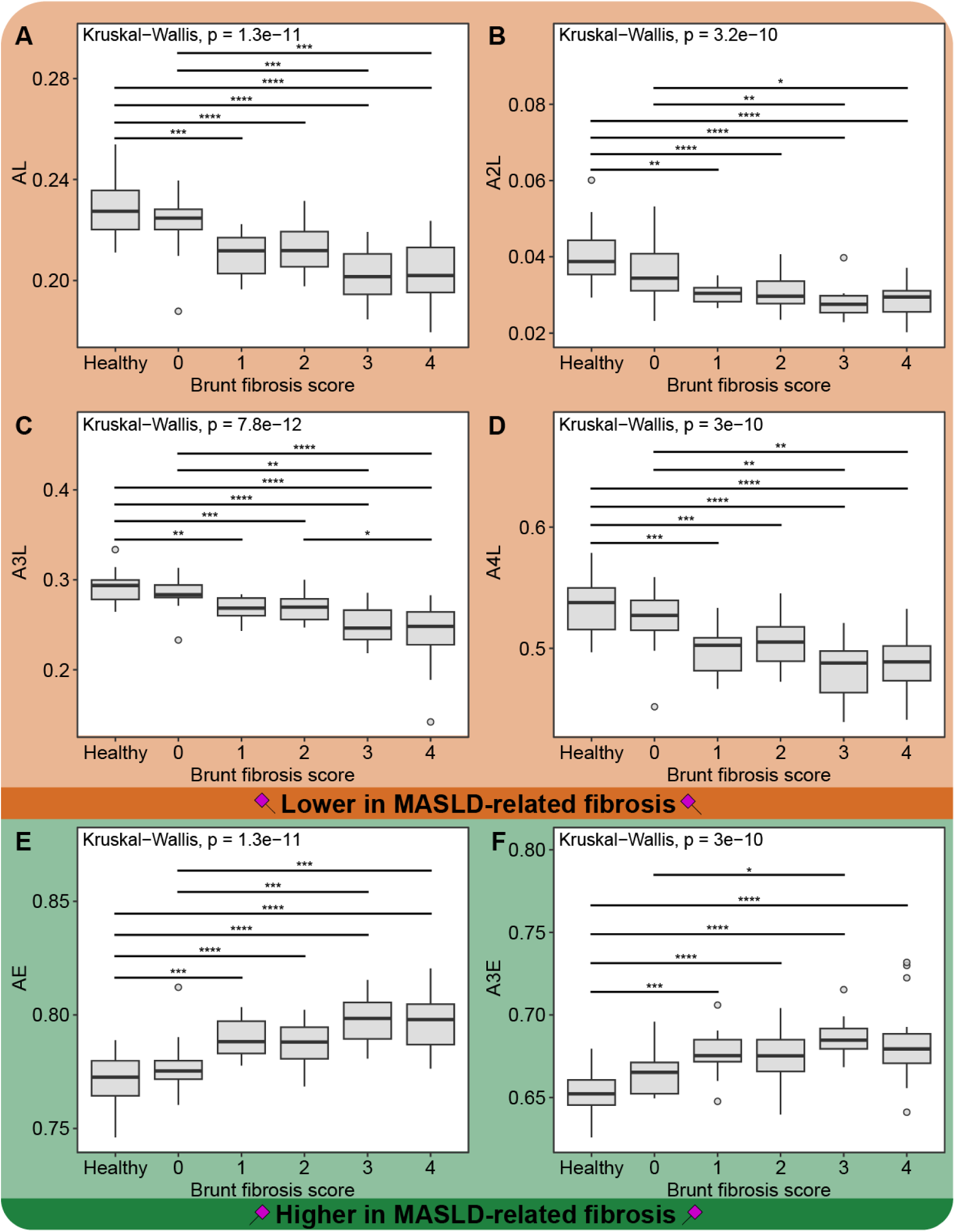
Replicated associations identified between patients with MASLD and healthy controls as stratified per MASLD-related fibrosis score (Brunt). *P*-values and fold changes are shown in **Supplementary Table 7-8**. *, **, ***: *p*-value < 0.05, 0.01, 0.001, respectively. The shown data corresponds to the replication cohort.

**Figure 4.**
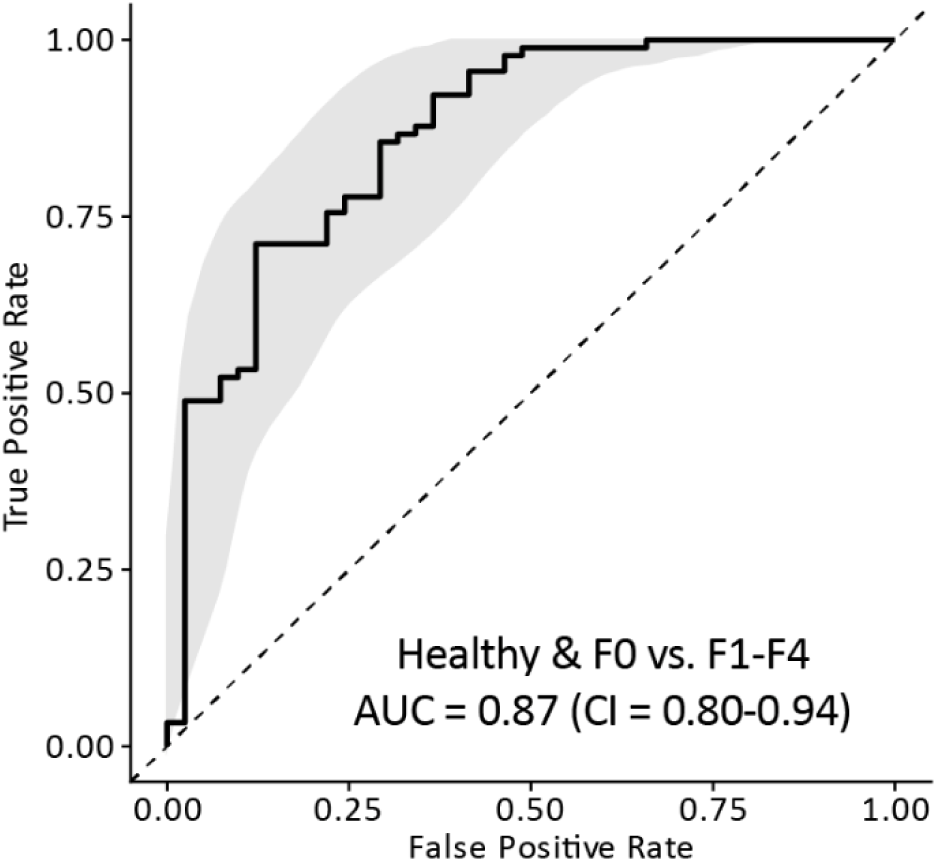
ROC analysis and associated AUC value for the single predictor model including A4L, illustrating its power to predict fibrosis upon histological appearance. Healthy controls and F0 patients were combined for this analysis.

**Figure 5.**
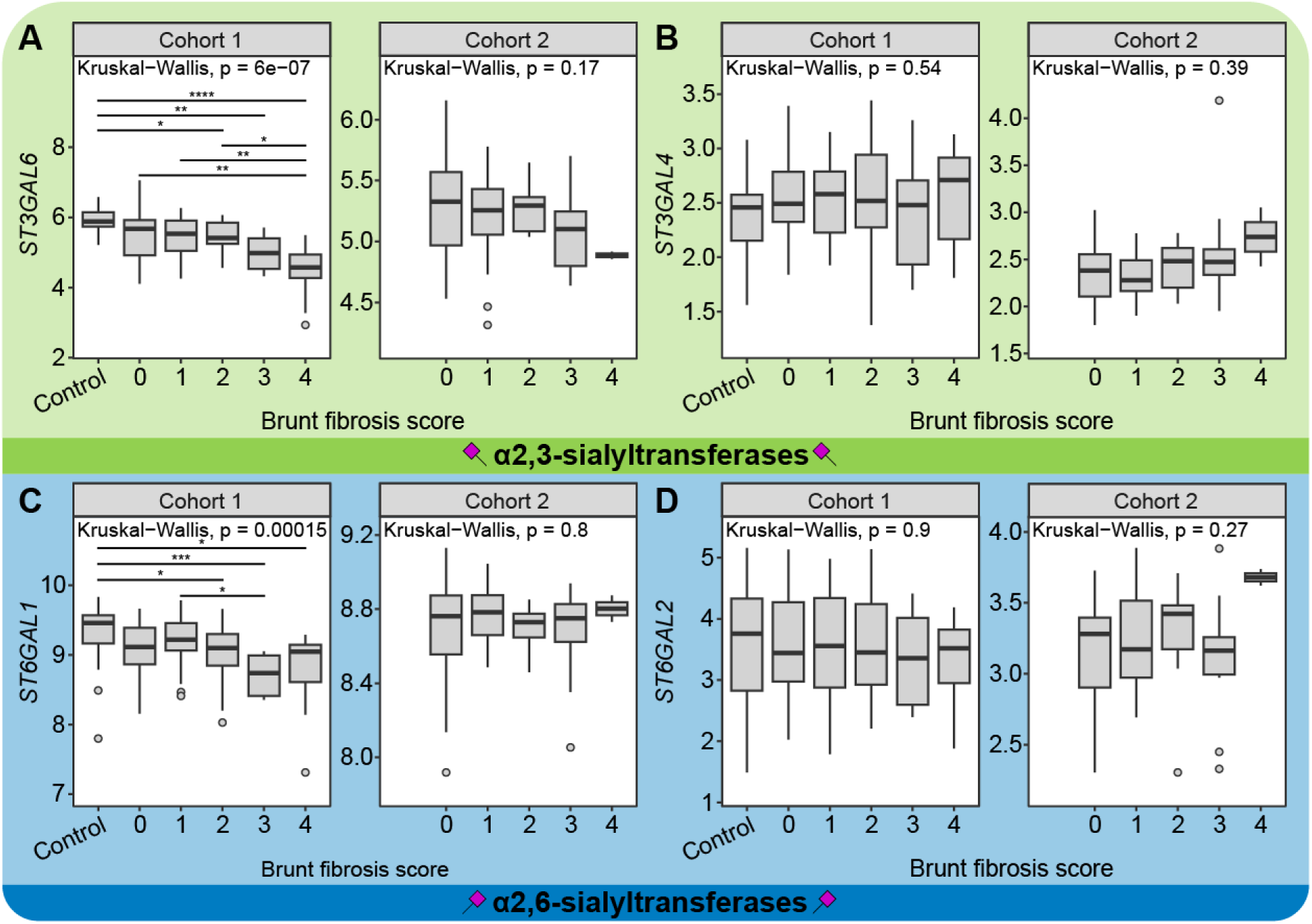
Associations identified between patients with fibrotic MASLD and controls with normal histology as stratified per MASLD-related fibrosis score (Brunt). *P*-values and fold changes are shown in **Supplementary Table 11-12**. *, **, ***: *p*-value < 0.05, 0.01, 0.001, respectively.

**Table 2.** Associations between blood *N*-glycan traits and MASLD as compared to healthy controls. Logistic regression was performed between MASLD (1) and healthy controls (0), including age, sex and their interaction as co-variates. Only replicated associations are shown. The results of all tests can be found in **Supplementary** Table 6. To account for multiple testing, *p*-values in the discovery cohort were corrected by the Benjamini-Hochberg procedure using a 5% false discovery rate.

| Derived traits | Description | *p*-value* |  | Odds ratio [95% CI] |  | Beta coefficient |  | Standard error |  |
| --- | --- | --- | --- | --- | --- | --- | --- | --- | --- |
|  |  | Disc. | Rep. | Disc. | Rep. | Disc. | Rep. | Disc. | Rep. |
| α2,6-linked sialylation |  |  |  |  |  |  |  |  |  |
| AE | Of A1-4 combined | 0.019 | 4.72 × 10<sup>-07</sup> | 2.75 [1.5 - 5.58] | 9.07 [4.19-23.71] | 1.01 | 2.20 | 0.33 | 0.44 |
| A3E | Of A3 | 0.019 | 4.33 × 10<sup>-07</sup> | 2.95 [1.61 - 6.04] | 11.6 [4.97-34.08] | 1.08 | 2.45 | 0.33 | 0.48 |
| α2,3-linked sialylation |  |  |  |  |  |  |  |  |  |
| AL | Of A1-4 combined | 0.019 | 4.72 × 10<sup>-07</sup> | 0.36 [0.18 - 0.67] | 0.11 [0.04-0.24] | -2.20 | -1.12 | 0.33 | 0.44 |
| A2L | Of A2 | 0.019 | 3.85 × 10<sup>-07</sup> | 0.3 [0.13 - 0.62] | 0.18 [0.09-0.33] | -1.70 | -1.61 | 0.40 | 0.33 |
| A3L | Of A3 | 0.019 | 1.53 × 10<sup>-06</sup> | 0.38 [0.19 - 0.7] | 0.1 [0.04-0.23] | -2.30 | -2.44 | 0.33 | 0.48 |
| A4L | Of A4 | 0.035 | 8.12 × 10<sup>-07</sup> | 0.45 [0.23 - 0.78] | 0.14 [0.06-0.28] | -1.98 | -2.52 | 0.30 | 0.40 |

CI: confidence interval.

**Table 3.** Correlation of conventional diagnostic markers, glycosylation signatures and glycosyltransferase expression levels with Brunt fibrosis score. Shown are the Spearman’s correlation coefficients (RS). Healthy controls were considered as F0 and pooled with patients with normal histology (F0) for this analysis.

| Variable | $R_s^{\#}$ | $p\text{-value}^{\#}$ |
| --- | --- | --- |
| NFS | 0.67 | $9.8 \times 10^{-14}$ |
| FIB4 | 0.66 | $1.4 \times 10^{-13}$ |
| A2L | -0.58 | $4.5 \times 10^{-13}$ |
| A3L | -0.66 | $8.2 \times 10^{-18}$ |
| A4L | -0.58 | $2.8 \times 10^{-13}$ |
| A3E | 0.55 | $1.7 \times 10^{-11}$ |
| AL/AE | -0.62 | $2.1 \times 10^{-15}$ |
| ST3GAL6 | -0.41 / -0.23 | $4 \times 10^{-7}$ / 0.045 |
| ST3GAL4 | 0.09 / 0.15 | 0.31 / 0.19 |
| ST6GAL1 | -0.30 / -0.03 | $3 \times 10^{-4}$ / 0.81 |
| ST6GAL2 | -0.08 / 0.06 | 0.36 / 0.61 |

<sup>#</sup>Cohort 1 / Cohort 2, respectively.

To evaluate the glycosylation traits, both individually and in combination, as predictors of fibrosis, we used logistic regression models with fibrosis status as the response variable. However, only the models with A3L and A4L as predictors passed rigorous model diagnostics and assumptions tests. Furthermore, due to strong collinearity between the predictors, the authors decided to use single-predictor models **(Supplementary** Figure 7), specifically AL4. **Figure 5** shows a Receiver Operating Characteristic (ROC) curve with CI space for the resulting model; the model metrics are summarized in the figure caption.

## Results

The blood protein *N*-glycome was analyzed by mass spectrometry, resulting in the identification of 81 and 54 *N*-glycans for the discovery and replication cohort, respectively. All *N*-glycans found in the discovery cohort were shared with those in the replication cohort, the lower number of identifications stems from more stringent data quality control settings in the latter. The annotated glycoforms were relatively quantified **(Supplementary Table 1, 2)** and summarized as glycosylation traits based on their structural features including fucosylation, antennary fucosylation, bisection, galactosylation, sialylation, antennarity and *N*-glycan type **(Figure 1, Supplementary Table 3, 4)**. The identified glycoforms were consistent with those commonly found on blood proteins, although the structures are putative and could include collections of isomers^46, 50^.

Blood protein *N*-glycosylation associates with MASLD

In a first step, we aimed to explore the differences between MASLD and healthy controls by principle component analysis, indicating that the separation between the two groups is driven by linkage-specific sialylation effects **(Supplementary** Figure 1). Further analysis revealed 6 replicated glycosylation traits between MASLD and healthy controls **(Figure 2**, **Table 2, Supplementary Table 6)**. These replicated glycosylation traits could indeed be classified into two major categories, namely α2,3- sialylated and α2,6-sialylated species **(Figure 2 b-g**, **Table 2)**.

Interestingly, lower α2,3-linked sialylation was observed across most complex-type *N*- glycans in patients with MASLD, regardless of antennarity. Specifically, the combined level of α2,3-sialylation over all complex-type glycans (AL) showed odds ratios of 0.36 and 0.11 for discovery and replication cohort, respectively **(Figure 2 b-e**, **Table 2)**.

In contrast, α2,6-sialylation (AE) was found to be generally increased in MASLD. This was most pronounced for *N*-glycans with three antennae (A3E; ORs of 2.95 and 11.6 in the discovery and replication cohort, respectively) **(Figure 2 f, g)**. This apparent shift toward lower α2,3-sialylation in MASDL was likewise conveyed by the AL/AE ratio **(Supplementary** Figure 2).

### Shifted linkage-specific sialylation status hallmarks fibrotic MASLD

As changes in α2,3- and α2,6-sialylation appeared to be a feature of MASLD, we further investigated these glycosylation traits in relation to the degree of fibrosis in patients enrolled in the replication cohort **(Figure 3, Supplementary Table 7-8)**. Patients without fibrotic scarring (non-fibrotic MASLD; Brunt fibrosis score 0) did not differ in the degree of their α2,3- **(Figure 4 a-d)** and α2,6-linked sialylation **(Figure 4 e, f)** from healthy individuals **(Supplementary Table 8)**. On the other hand, linkage-specific sialylation effects were observed upon the histological manifestation of fibrosis (MASLD with various degrees of fibrosis; Brunt fibrosis score 1-4) **(Supplementary Table 8)**. Importantly, these associations were neither affected by demographics (age, sex, BMI) nor by the common comorbidity, T2DM status, of the individuals **(Supplementary** Figure 3**, 4, 7, Supplementary Table 11)**. Furthermore, no association was found between levels of overall sialylation (that is, without the distinction of linkage isomers) and early fibrosis stages **(Supplementary** Figure 5). Additionally, we performed a similar analysis including the conventional non-invasive diagnostic markers of fibrosis, FIB-4 and NFS. In line with literature-based expectations^51^, these markers appeared to be best at differentiating advanced, but not early, fibrosis from non-fibrotic conditions **(Supplementary** Figure 6).

Motivated by the found associations, we conducted a ROC analysis to evaluate the discriminative power of the glycosylation traits to predict the histological manifestation of fibrosis. Using a “step” function, we obtained an optimal model with A4L as a single predictor (OR: 0.13, CI = 0.06-0.26, *p*-value < 0.0001), of which associated ROC curve is shown in **Figure 4** with CI space. Additional model parameters were: AUC = 0.87, CI = 0.80-0.94. Similar results were obtained for A3L as a single predictor, suggesting that the best-performing predictors of fibrotic MASLD are A4L and A3L. The inclusion of potential confounding factors such as age, sex, BMI and T2DM status did not improve these models. Due to limited sample numbers in the F0 group (n=12), the same analysis could not be performed for the conventional diagnostic markers FIB-4 and NFS.

Fibrosis is not the single determinant of MASLD-MASH severity. Another important histological scoring system is NAS, used to assess the severity of liver inflammation and damage in patients with MASLD-MASH as well as its progression propensity. This composite score includes three components: steatosis (fat accumulation), lobular inflammation (presence of inflammatory cells) and ballooning (swelling/damage) of liver cells. Except for some weak correlations, none of these associated with sialylation and the blood *N*-glycome in the current study **(Supplementary** Figure 7).

### Liver sialyltransferase expression patterns associate with fibrotic MASLD

Little is known about the regulation of glycosylation in the liver. The observed alterations with fibrosis can have different origins, for example being driven by the up- or downregulation of glycan carrying proteins or receptors regulating their half-life, by the availability of sialic acid donors, or by the altered expression of sialyltransferases (i.e., the enzymes responsible for sialylating glycoproteins). To investigate the latter, two publicly available total RNA-seq datasets were re-analyzed^48, 49^, with focus on *N*- glycan-specific sialyltransferase (α2,3-sialylation: *ST3GAL4*, *ST3GAL6*; α2,6-sialylation: *ST6GAL1*, *ST6GAL2*)^52^ transcript expression in liver tissue from individuals with normal histology and patients with (fibrotic) MASLD. We found the expression levels of *ST3GAL6* to follow a decreasing trend with fibrosis **(Figure 5a**, **Table 3)**, displaying noticeable similarities to the observed glycomics pattern with regards to α2,3-sialylation **(Figure 3a-d)**, whilst no changes were observed for *ST3GAL4* **(Figure 5b**, **Table 3)**. Later stage fibrotic biopsies showed a lower *ST6GAL1* expression relative to controls only in Cohort 1 **(Figure 5c**, **Table 3)**, whereas no strong effects were found for the expression of *ST6GAL2* **(Figure 5d**, **Table 3)**. It could be hypothesized that our observations on the globally lower α2,3-linked sialylation of *N*- glycans might be explained by the direct regulation of this type of sialylation via *ST3GAL6* downregulation. The more specific increase of α2,6-linked sialylation on tri-antennary *N*-glycans is considered to rather be a protein-specific effect.

### Correlation of conventional diagnostic, glycomic and transcriptomic signatures with fibrosis stage

To further investigate the glycomic associations found in the study, an exploratory ranked correlation analysis was performed between conventional diagnostic markers, replicated glycosylation traits as well as sialyltransferase expression levels versus Brunt fibrosis scores, with the aim to evaluate the potential of these markers for staging fibrosis. Using this approach, we found both the glycomic markers and conventional diagnostics markers (FIB-4 and NFS) as strong correlators with fibrosis grade **(Table 3)**. However, while conventional markers seem to be effective in differentiating between advanced fibrosis stages, they do not reliably indicate early fibrotic development **(Supplementary** Figure 6). In contrast, reduced α2,3-sialylation seems to be a hallmark of fibrosis from its initial histological manifestation **(Figure 3, Supplementary** Figure 6). Similar observations were made for the AL/AE ratio, which as an overall descriptor of linkage-specific sialylation status also significantly correlated with the progression of fibrosis, suggesting that the found glycan markers follow a unidirectional trend and might be phenotype-indicative **(Table 3)**. Furthermore, we found the transcript levels of *ST3GAL6*, a key α2,3-sialyltransferase, to negatively correlate with fibrosis stage in both cohorts **(Table 3)**.

## Discussion

In the present study, we identified a characteristic, to our knowledge unreported, hallmark of (fibrotic) MASLD, as we show in two independent cohorts that this patient group displays globally lower α2,3-linked sialylation on their circulation proteins as compared to age- and sex-matched healthy individuals. Importantly, the aforementioned glycosylation signatures discerned fibrotic from non-fibrotic MASLD, with histology assessed by two independent pathologists, and appears to be unaffected by the common comorbidity T2DM.

While liver biopsy is the golden standard for diagnosis of fibrosis, this is not suitable as a screening tool in the general population. Noninvasive tests or a combination of selected noninvasive tests are increasingly used in hepatology practices, but so far lack diagnostic accuracy in low risk general populations to be used as a reliable screening test^15, 53^. Alterations in blood protein *N*-glycosylation patterns, as discovered here, can serve as non-invasive biomarkers of liver diseases as shown previously in the diagnosis of cirrhosis and hepatocellular carcinoma^17^.

Sialic acid linkage isomers exhibit functionally distinct roles, and changes in the abundance of one linkage variant over the other have been reported in cancer, inflammatory bowel disease and T2DM^47, 54–57^. Previously, we found decreased α2,3- sialylation in patients with T2DM, albeit together with other glycosylation signatures marking this condition^38, 47, 57^. Many patients with a metabolic disease develop MASLD over time, with a consequent propensity to progress to fibrotic MASLD-MASH^2^. Therefore, assessing the interference of T2DM on the reported glycomics signature is critical. Interestingly, our analysis on T2DM status stratified groups suggests that low α2,3-sialylation is fibrosis- and not T2DM-specific. While this finding requires validation, we hypothesize that low α2,3-sialylation in earlier T2DM glycomics studies may merely indicated fibrosis in the patients^47, 57^.

Regarding glycomics of (fibrotic) MASLD, larger-scale studies relying on sialic acid linkage isomer differentiation as reported in the current work have hitherto been lacking. Yet, a previous study of 15 patients across 3 fibrosis categories found a decrease in the ratio of non-fucosylated to fucosylated glycans that are fully sialylated and carry a single α2,3-linked sialic acid with fibrosis progression, concluding that their observed signatures are antennary fucosylation-dependent and likely originate from the acute phase protein alpha-1-antitrypsin ^27^. Another research group found a signature linked to the aforementioned composition (i.e., triantennary fully sialylated *N*-glycans) as being linked to both alpha-1-antitrypsin and alpha-1-acid-glycoprotein, although no details were reported on the type of fucosylation and sialic acid linkages^21^. Furthermore, composite markers such as the GlycoFibroTest^20^ and the more recent GlycoFibroTyper^25^ rely on total serum or affinity-enriched immunoglobulin G *N*- glycosylation analysis, and both suggest the presence of a bisecting *N*- acetylglucosamine (that is, bisection) as an important glycosylation feature differentiating clinical phenotypes of MASLD.

As portrayed above, glycomic studies have reported *N*-glycan signatures that associate with fibrosis, and directly^21^ or indirectly^20, 27^ related these glycans to candidate proteins^17^. Of note, changes in specific glycoprotein abundance may, to some extent, explain the observed relative glycoform frequency changes in our total blood analysis. Noticeably, our glycomics data points towards globally lower α2,3- sialylation, as the complete range of glycans, from di- to tetra-antennary species is affected. Additionally, no significant alterations in major plasma glycoprotein concentrations have been described by proteomics studies^58^, supporting our hypothesis that changes in the *N*-glycan biosynthetic pathway or glycan-dependent protein clearance are at the basis of the observed blood *N*-glycome fibrosis signatures. The first option is further substantiated by the reduced expression of a hepatic glycosyltransferase (*ST3GAL6*) responsible for the addition of sialic acids to *N*-glycans in α2,3-linkage, as we found in the re-analysis of publicly available transcriptomics dataset comparing the different stages of MASLD-related fibrosis. We hypothesize that this observation speaks for a potential disruption of sialylation in hepatocytes in the setting of liver fibrosis before glycoproteins are secreted into the circulation.

The half-life of glycoproteins in the blood is regulated by glycan-recognizing receptors, such as the asialoglycoprotein receptor (also known as Ashwell-Morell receptor) clearing non-sialylated glycoproteins from the circulation. As this receptor has a weak affinity for α2,6-linked sialic acids^59^, a decrease in the expression of the asialoglycoprotein receptor could result in an increase in α2,6-linked sialylation on blood proteins. Assessment of glycosyltransferase transcript expression levels of *ASGR1* and *ASGR2*, corresponding to asialoglycoprotein receptor subunits, did not conclusively support such a hypothesis (data not shown).

Interestingly, a study using a hepatocyte-specific conditional knockout of glycosyltransferase β-galactoside α2,6-sialyltransferase 1 (*St6gal1*; encoding the enzyme responsible for the addition of sialic acids in an α2,6-linkage) in mice connected the loss of hepatocyte and circulatory glycoprotein α2,6-sialylation to the spontaneous development of fatty liver disease and a shift towards a proinflammatory immune-phenotype^60^. In contrast, we found low α2,3-sialylation and elevated α2,6- sialylation on *N*-glycans in patients with MASLD-related fibrosis. As the latter feature was restricted to triantennary glycans in our data, it may be linked to an increase in the abundance of proteins carrying such glycoforms, including but not limited to previously reported acute phase proteins such as alpha-1-antitrypsin ^21, 61^. To further unravel this, it would be necessary to subject alpha-1-antitrypsin-associated glycans or glycopeptides for a sialic acid linkage-specific analysis.

While the molecular mechanisms shaping glycosylation in fibrotic MASLD-MASH remain to be elucidated, the striking similarity of blood protein *N*-glycosylation patterns in pancreatic cancer should not be overlooked^39, 62^. Cancer-associated fibroblasts play critical roles in extracellular matrix formation across various cancers, including pancreatic, lung and liver cancers^63^. However, their presence is a particularly notable pathological feature of pancreatic cancer, and has been reported to metabolically reprogram tumor and immune cells, thereby inducing a favorable microenvironment for fibrosis development and tumor progression^63, 64^. Such a fibrotic (tumor) microenvironment secretes a variety of paracrine mediators^63^ that may influence glycan biosynthetic pathways in subtle, currently unknown ways, and may lead to *ST3GAL6* downregulation in the liver, resulting in the here reported glyco-phenotype. Future glycomic studies should therefore consider fibroblast-positive cancers and other gastroenterological disorders (e.g. alcohol-, drug-, viral- or autoimmune-induced liver diseases) as a potential source of low blood protein α2,3-sialylation. Collectively, this could indicate that the described glycosylation signature is a universal fibrosis hallmark rather than a specific feature of fibrotic MASLD.

The current study has potential implications for the diagnosis of MASLD- related fibrosis, albeit this needs to be confirmed with a longitudinal, prospective study setup in a larger patient population. Moreover, while certain glycosylation traits exhibit sex- and age-specific alterations, the glycome in healthy individuals at a certain age is relatively stable and well predictable^39^, allowing to establish diagnostic benchmark levels. Nevertheless, the employed technology needs further development to reach clinical application. At present, the clinically available GlycoFibroTest and GlycoFibroTyper do not include (linkage-specific) sialylation analysis^20,25^. Therefore, alternative glycoanalytical platforms employing lectin-affinity towards specific sialic acid linkage isomers, or separation techniques capable of resolving multi-antennary isomeric glycans may better aid clinical translation^65–68^.

## Conclusions

In conclusion, a replicated fibrosis-specific blood *N*-glycosylation signature was found in MASLD, which allows the detection of fibrosis early-on. The global decrease of α2,3- sialylation, a unique feature of circulatory proteins produced by the fibrotic liver, offers the possibility for developing novel non-invasive diagnostic tests that facilitate early diagnosis. The possibility of analyzing dried blood spots can be a significant advantage for use as a general screening tool^44^. Furthermore, the observed glycomic and transcriptomic signatures point toward molecular mechanisms that may play a role in the development of fibrosis. Clinical translation of the glycomic signature has the prospects of making diagnosis and monitoring of fibrosis more comfortable for both patients with MASLD and physicians in the future and may allow for timely intervention and improved disease management.

## Data Availability

Raw mass spectrometry data will be made available upon reasonable request via a designated public repository. Analytical methods are described in sufficient detail in the manuscript and supplementary methods, but further information will be provided upon request. Study material, i.e., human blood samples, will not be made available.

## Acknowledgements

We express our gratitude to dr. Marta Kanabus for her invaluable insight that influenced the genesis of this work.

## Transcript profiling

Re-analysis of GSE130970 (https://www.ncbi.nlm.nih.gov/geo/query/acc.cgi?acc=GSE130970) and GSE162694 (https://www.ncbi.nlm.nih.gov/geo/query/acc.cgi?acc=GSE162694).

## Authors’ Contributions

T.P.: Study design, sample preparation, data curation, formal analysis, validation, investigation, visualization, statistical analysis, data interpretation, writing – original draft preparation. B.V.: Study design, data acquisition, sample collection, ethical application, writing – editing. A.L.M: sample collection, data acquisition, writing – editing. O.M.: Data curation, visualization, statistical analysis. S.N.: data acquisition. M.R.B.: sample preparation. W.W.: Sample preparation. M.B.: Study design. M.N.: writing – editing, study design, funding acquisition. J.V.: writing – editing, data analysis. A.G.H.: Sample collection, data acquisition, writing – editing. B. van H.: Supervision, funding acquisition, writing – editing. N. de H.: Study design, data interpretation, writing – editing. M.W.: Supervision, data interpretation, funding acquisition, writing – editing. M.T.: Supervision, study design, data interpretation, writing – original draft preparation, writing – editing, funding acquisition.

All authors were involved in the critical revision of the manuscript and have approved the final version of the manuscript.

## Abbreviations

MALDI-FTICR-MS: Matrix Assisted Laser Desorption/Ionization - Fourier Transform Ion Cyclotron Resonance – Mass Spectrometry
MASLD: Metabolic dysfunction-associated steatotic liver disease
MASH: Metabolic dysfunction-associated steatohepatitis
T2DM: Type 2 Diabetes Mellitus
ROC: Receiver Operating Characteristic

