## Supplementary Methods and Figures for "Blood *N*-glycomic signature of fibrosis in MASLD shows low levels of global α2,3-sialylation"

### SUPPLEMENTARY MATERIAL

#### Blood *N*-glycomic signature of fibrosis in metabolic-dysfunction associated steatotic liver disease shows low level of global α2,3-sialylation

Tamas Pongracz[1,5], Bart Verwer[2], Anne Linde Mak[3], Oleg A. Mayboroda[1], Simone Nicolardi[1], Marco René Bladergroen[1], Wenjun Wang[1], Maaike Biewenga[2], Max Nieuwdorp[3], Joanne Verheij[4], A.G. (Onno) Holleboom[3], Bart van Hoek[2], Noortje de Haan[1], Manfred Wuhrer[1,*], Maarten E. Tushuizen[2,*]

[1]Center for Proteomics and Metabolomics, Leiden University Medical Center, The Netherlands

[2]Department of Gastroenterology and Hepatology, Leiden University Medical Center, The Netherlands

[3]Department of Internal Medicine, Amsterdam University Medical Center, Amsterdam, The Netherlands

[4]Department of Pathology, Amsterdam University Medical Center, Amsterdam, The Netherlands

[5]Present address: Department of Clinical Sciences, Karolinska Institutet Danderyd Hospital, Stockholm, Sweden

**Supplementary Materials & methods**

*Materials*

Materials and reagents used in the study were of analytical grade and purchased from commercial suppliers. Type I Ultrapure Water (UP) was used to prepare solutions, which was produced by an ELGA Purelab Ultra system (Elga LabWater, High Wycombe, United Kingdom). Nonidet P-40 substitute (NP-40), super-DHB and 1-hydroxybenzotriazole monohydrate (HOBt), ammonium bicarbonate ($NH_4HCO_3$), potassium chloride (KCl), disodium hydrogen phosphate hydrate ($Na_2HPO_4{\cdot}7H_2O$) and 85% phosphoric acid ($H_3PO_4$) were obtained from Sigma-Aldrich (Steinheim, Germany). Ethanol, sodium hydroxide (NaOH), sodium dodecyl sulphate (SDS), and trifluoroacetic acid, disodium hydrogen phosphate dihydrate ($Na_2HPO_4{\cdot}2H_2O$), potassium dihydrogen phosphate ($KH_2PO_4$), and sodium chloride (NaCl) were purchased from Merck (Darmstadt, Germany). 1-ethyl-3-(3-(dimethylamino)propyl)carbodiimide hydrochloride (EDC) was obtained from Fluorochem (Hadfield, United Kingdom), while peptide-*N*-glycosidase F (PNGase F) was purchased from Roche Diagnostics (Mannheim, Germany). HPLC-supra-gradient acetonitrile (ACN) and ethanol (EtOH) were obtained from Biosolve (Valkenswaard, The Netherlands) and Merck (Darmstadt, Germany), respectively. The Visucon-F healthy human plasma standard originated from Affinity Biologicals (Ancaster, Canada). Peptide Calibration Mix II was obtained from Bruker Daltonics (Billerica, MA).

*Sample preparation for matrix-assisted laser desorption/ionization – Fourier transform ion cyclotron resonance – mass spectrometry (MALDI-FTICR-MS)-based high-throughput glycosylation analysis*

Plasma samples of the discovery cohort were part of a larger preceding study on autoimmune hepatitis, and were randomized on overall 5 96-well plates, together with 4 Visucon F standards and 2 or 3 blanks per plate[1]. Serum samples of the replication study were randomized on a single 96-well plate, together with 4 pools (i.e. a pool generated by pooling equal amounts of serum from each patient in the cohort). Age and sex were taken into account for an optimal distribution of cases and controls per plate.

*Glycan release from sera/plasma, linkage-specific sialic acid stabilization and MALDI-FTICR-MS analysis*

Release of *N*-glycans from plasma proteins and linkage-specific chemical sialic acid derivatization was performed as previously described in similar high-throughput, robotized workflow, using 2 uL of plasma/serum for the release[2, 3]. For MALDI-FTICR-MS measurement, 1 uL sDHB matrix was topped by 1uL HILIC-purified sample and left to dry by air[2, 3]. The measurement was performed on a 15T Bruker SolariX XR FTICR mass spectrometer equipped with a ParaCell, a Smartbeam-II laser and a Combisource (Bruker Daltonics, Bremen, Germany) in positive ionization mode[3]. Prior to measurement, calibration was performed with Peptide Calibration Mix II (Bruker Daltonics). For each spot, an average spectrum was obtained from the acquisition of 10 spectra in the m/z range of 1000–5000 using 1 M data points.

*Data processing*

MALDI-FTICR-MS raw spectra were converted into xy files. Extraction of these raw data was performed using in-house developed software MassyTools[4]. For the targeted extraction of glycan peaks, analyte lists were created based on manual annotation of summed mass spectra. The assignment of glycoforms was based on exact mass and previous reports[3, 5-7]. The $1^+$ charge state was used for extraction. Signals were integrated by covering minimum 95% of the area of the isotopic envelope of glycan peaks. An analyte was included in the final data analysis if its signal-to-noise was above 27, its isotopic pattern did not deviate more than 25% from the theoretical one, and if its mass error was within a ±20 parts per million range. Additionally, the same analyte had to be present in at least 1 out of 4 spectra (25%) in spectra per disease group for inclusion to the final data analysis. The relative intensity values of glycan compositions that passed quality criteria were calculated by normalizing to the sum of their total areas.

*Glycosylation trait calculation*

Based on the measured blood-derived glycan traits (n = 81 and 72 for the discovery and replication cohort, respectively **(Supplementary Table 1, 2)**), glycosylation traits were calculated based on common structural characteristics, including the number of antennae (A), and the levels of fucosylation (F), antennary fucosylation (Fa) bisection (B), galactosylation (G), or sialylation (S), **(Figure 1, Supplementary Table 3-4)**.

*Method repeatability and robustness*

To assess the MALDI-FTICR-MS method repeatability, the inter-plate coefficient of variation of the most abundant glycan peak H5N4E2 was calculated for the plasma standards in the discovery cohort and the intra-plate variation of the pools for the same glycan in the replication cohort and were 4.9% and 1.2%, respectively.

*Statistics*

Age, sex and their interaction were included as co-variates in a logistic regression model to find disease specific associations of HC (0) versus MASLD (1). The odds ratios (OR) were calculated with their 95% confidence intervals (CI) and represent single standard deviation increases in the tested derived traits. Multiple testing correction was performed using the Benjamini-Hochberg procedure and was based on a false discovery rate (FDR) of 5% in the discovery cohort. On the analogy of genome wide association studies, we used the discovery cohort to identify potential glycomic associations between healthy individuals and those with MASLD. In the discovery study, multiple testing correction was applied to avoid/limit the finding of false positive associations. In contrast, the replication cohort was used to confirm associations previously found in the discovery cohort. To avoid overlooking valid associations, we found it less necessary to perform multiple testing correction in the replication cohort. The statistical analyses were performed in R, version 4.2.2 (R Foundation for Statistical Computing, Vienna, Austria) and RStudio, version 2022.12.0, Build 353 (RStudio, Boston, MA).

#### Supplementary Figures

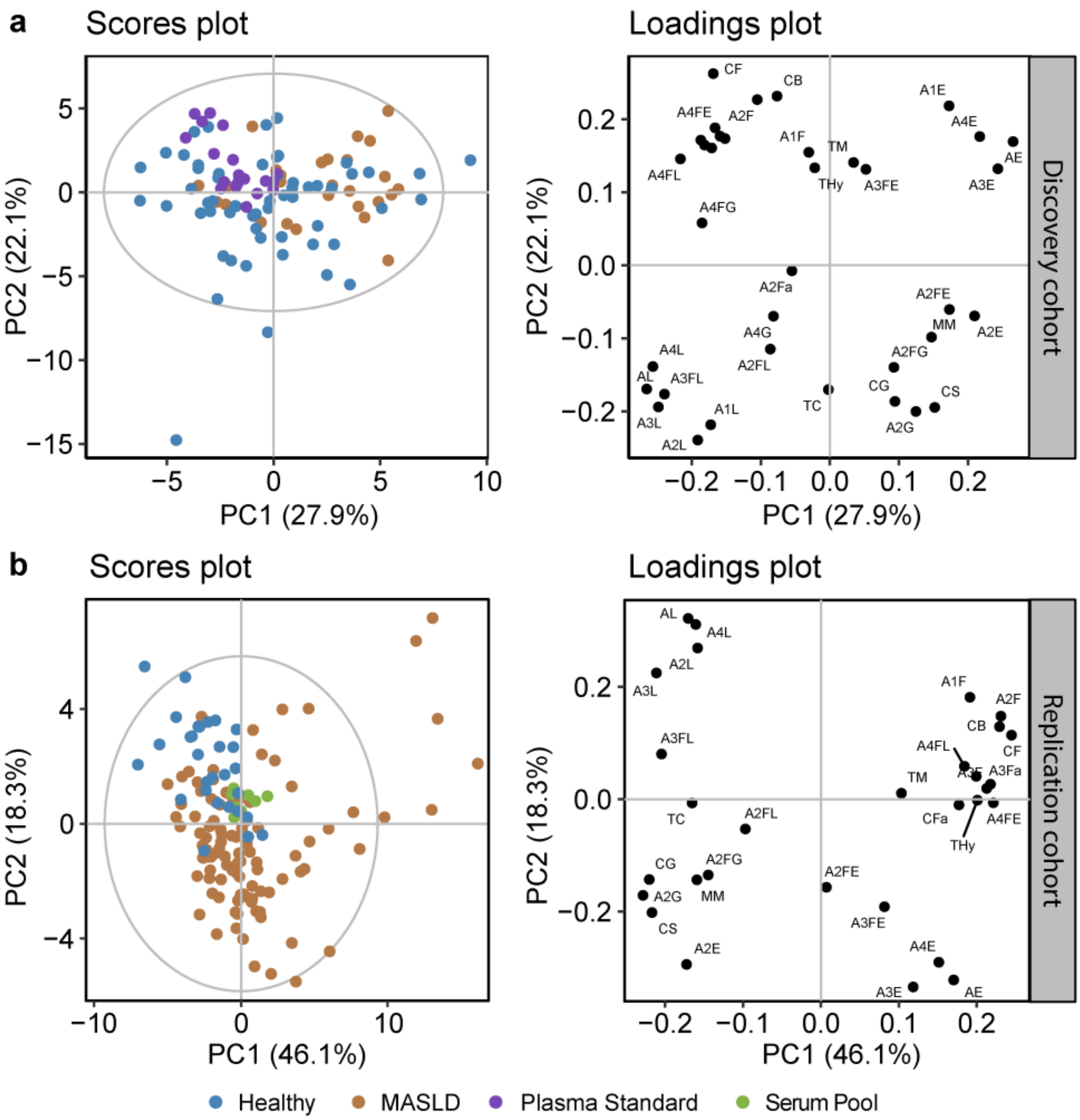

**Supplementary Figure 1. Principle component analysis (PCA) based on the calculated glycosylation traits in each cohort, illustrating the distribution of samples and standards or pools.** Scores plot of the PCA analysis for the discovery **(a)** and replication **(b)** illustrating the separation between healthy controls (blue) and MASLD patients (purple) along PC1 and PC2, as well as the clustering of standards (discovery cohort, **a**) or pools (replication cohort, **b**). Loadings plots visually represent the variables that contribute to the separation in the PCA model in each cohort. Plasma standards in the discovery cohort **(a)** were distributed across 5 plates, and have previously been shown to be randomly scattered, indicating the absence of systematic batch effects[1]. The close clustering of plasma standards in the discovery cohort and pools in the replication cohort indicate the low technical variability of the method.

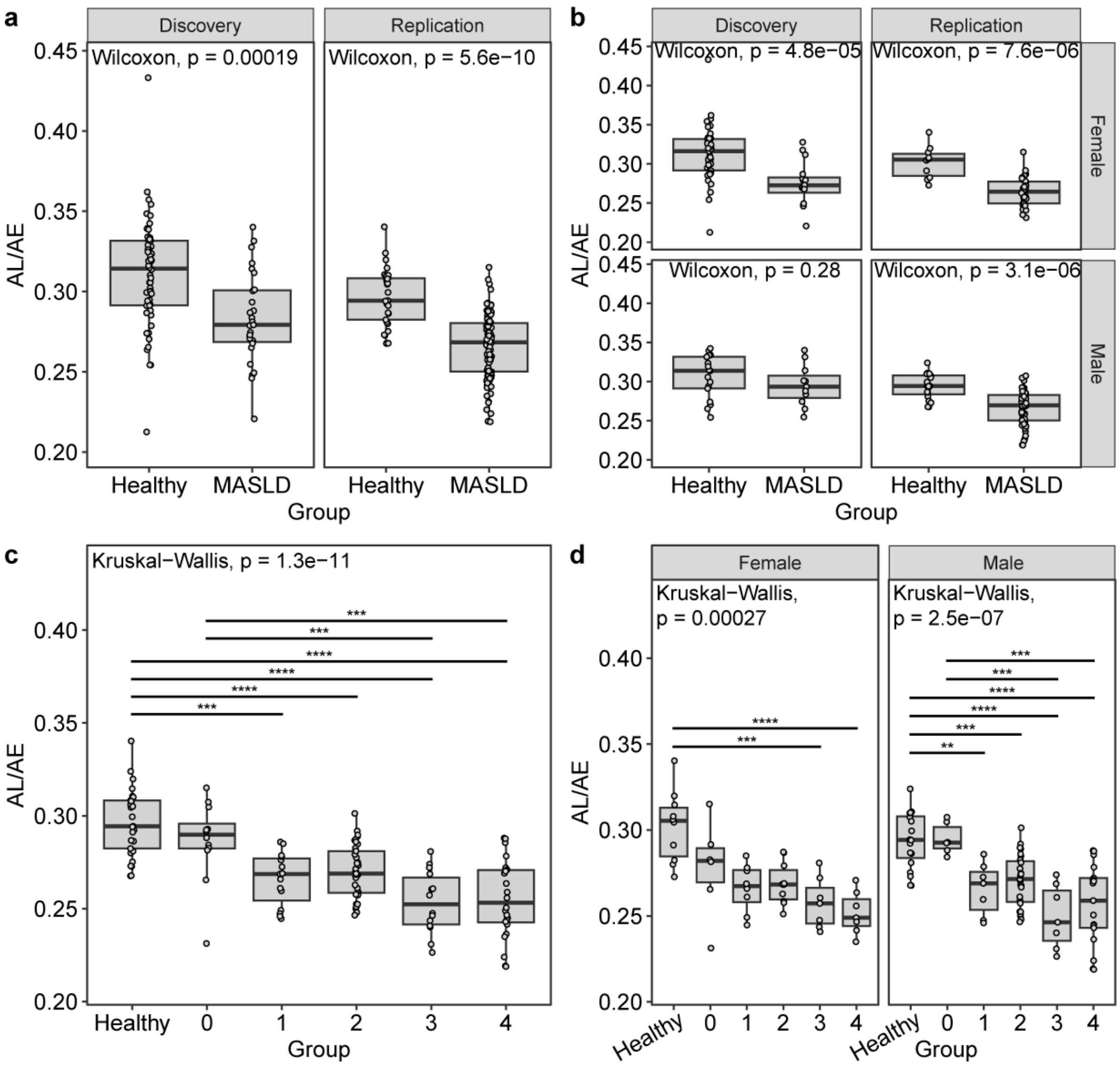

**Supplementary Figure 2. The ratio of the glycosylation traits AL and AE, illustrating the observed effect of NAFLD (a-b), and more specifically of fibrosis (c-d) on the blood *N*-glycome as a single glycosylation trait. (a)** Differences in the ratio of AL/AE in healthy controls versus NAFLD patients overall, and **(b)** sex stratified in the cohorts. **(c)** Overall and **(d)** sex stratified differences in AL/AE between healthy controls and patients with various degree of fibrosis (Brunt fibrosis score) in the replication cohort. While this ratio represents the overall effect, due to its limitations described in the results and discussion, all significantly associated glycosylation traits are shown and described separately in the main manuscript **(Figure 2, 3; Table 2, 3)**. Note that sex stratification results in lower power for statistical analysis, especially for females **(d)**. As age and sex is rather well-matched between the groups, we believe that the less differentiation between healthy & F0 vs. F1-F4 fibrosis scores for the female group is due to low sample size. Furthermore, neither sex nor age were founds as significant covariates in binary classification models.

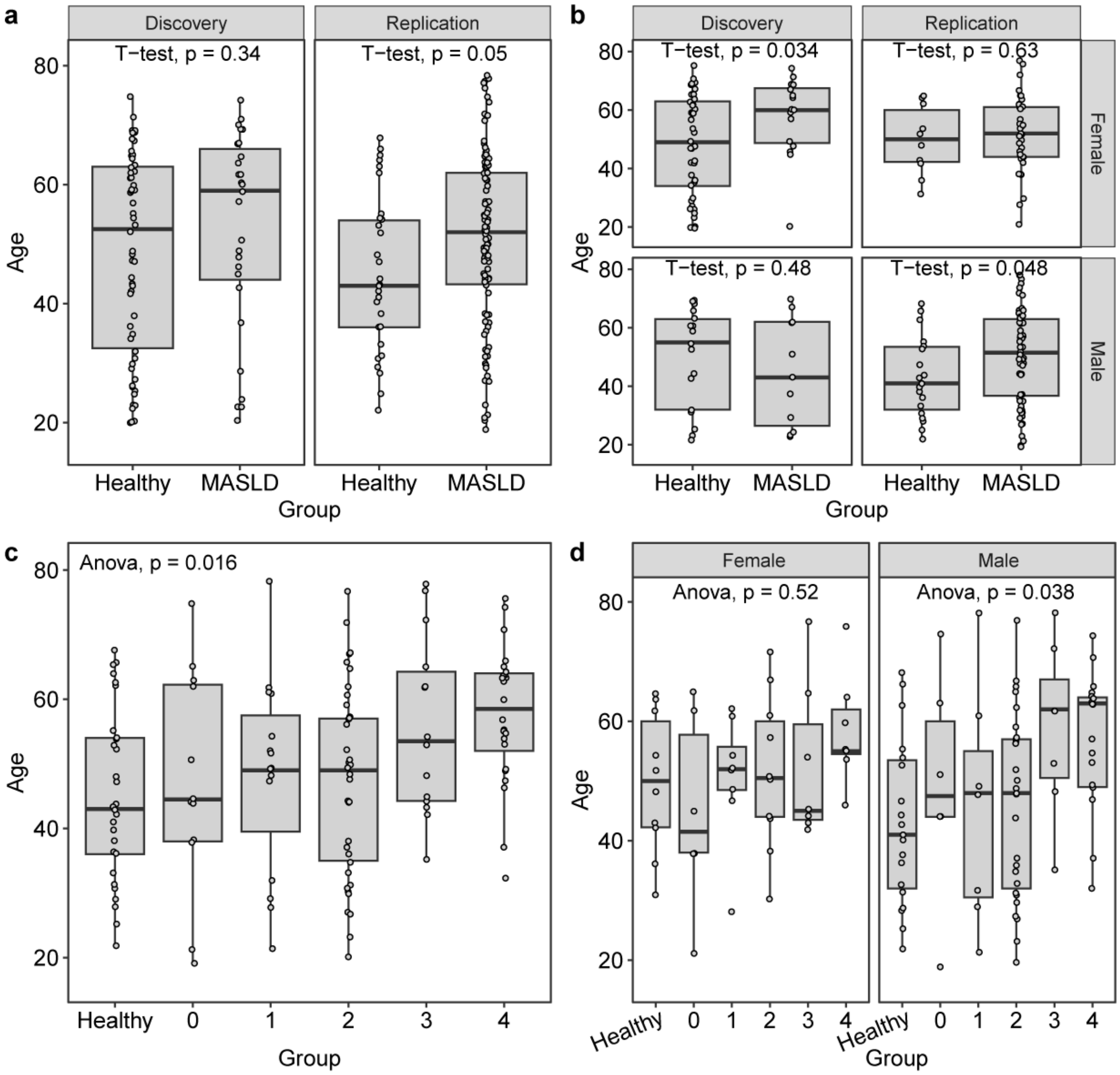

**Supplementary Figure 3. Age and sex distribution across disease groups (a)** Overall and **(b)** sex stratified age distribution in the cohorts. **(c)** Overall and **(d)** sex stratified age distribution in the replication cohort. A significant age difference was observed for females in the discovery cohort and for males in the replication cohort based on simple statistical testing, although this effect has been accounted for by logistic regression analysis including age, sex and their interaction in the model as covariates **(see "Statistics" section of the Supplementary Material)**. Furthermore, neither sex nor age were found as significant covariates in binary classification models.

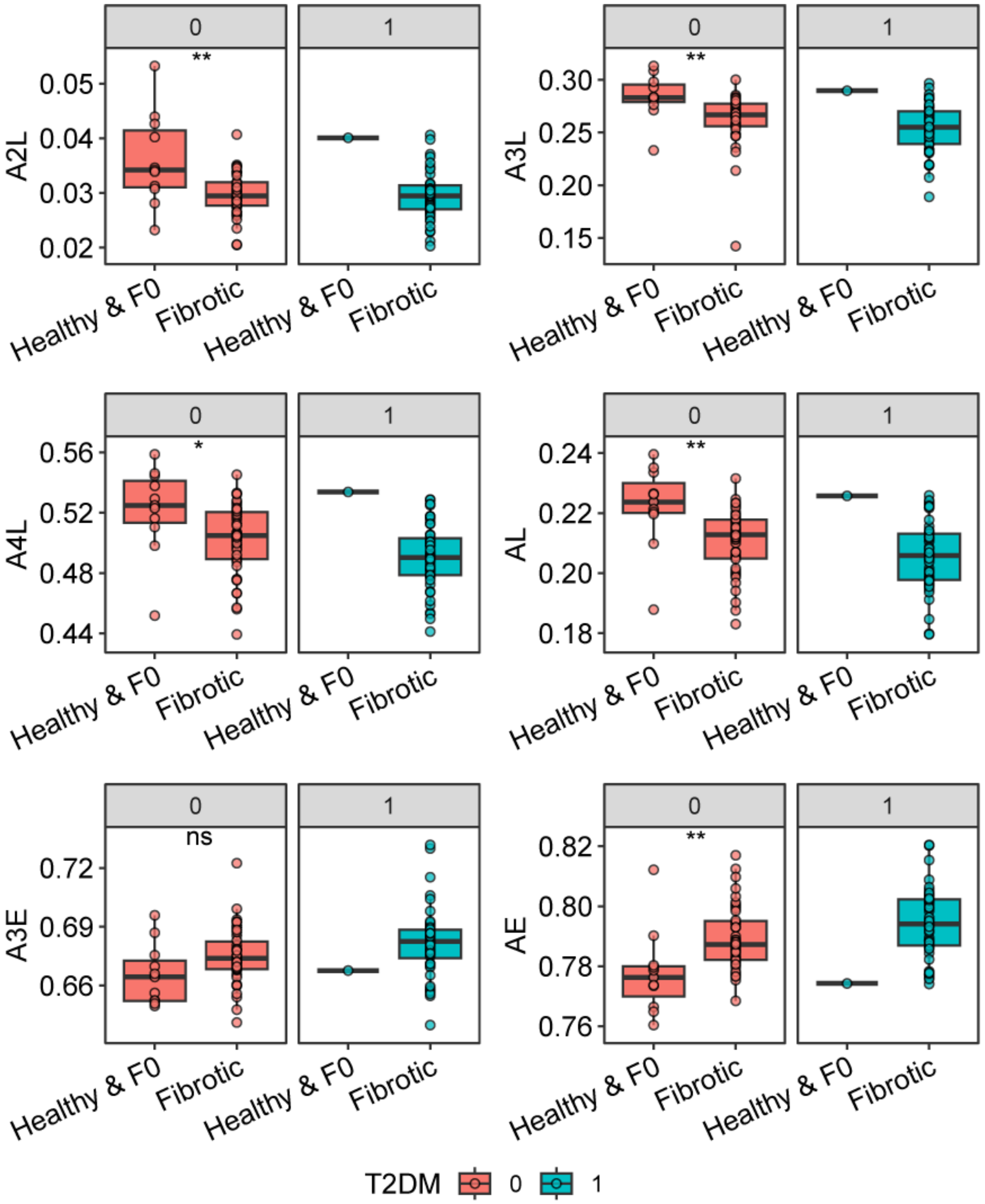

**Supplementary Figure 4. Influence of type 2 diabetes mellitus (T2DM).** Association of glycosylation signatures and fibrosis in T2DM stratified groups (0 (red) = no T2DM; 1 (blue) = T2DM). Healthy patients were considered F0 and non-diabetic. For the applicable groups, healthy and F0 patients were pooled for this analysis. Note that due to the low number of F0 patients in the T2DM group, and low number of patients upon further stratification per fibrosis stage, further univariate statistical analysis could not be performed. However, the direction of alteration upon fibrosis is similar in both the non-T2DM and T2DM groups, indicating a fibrosis- and not T2DM-specific effect.

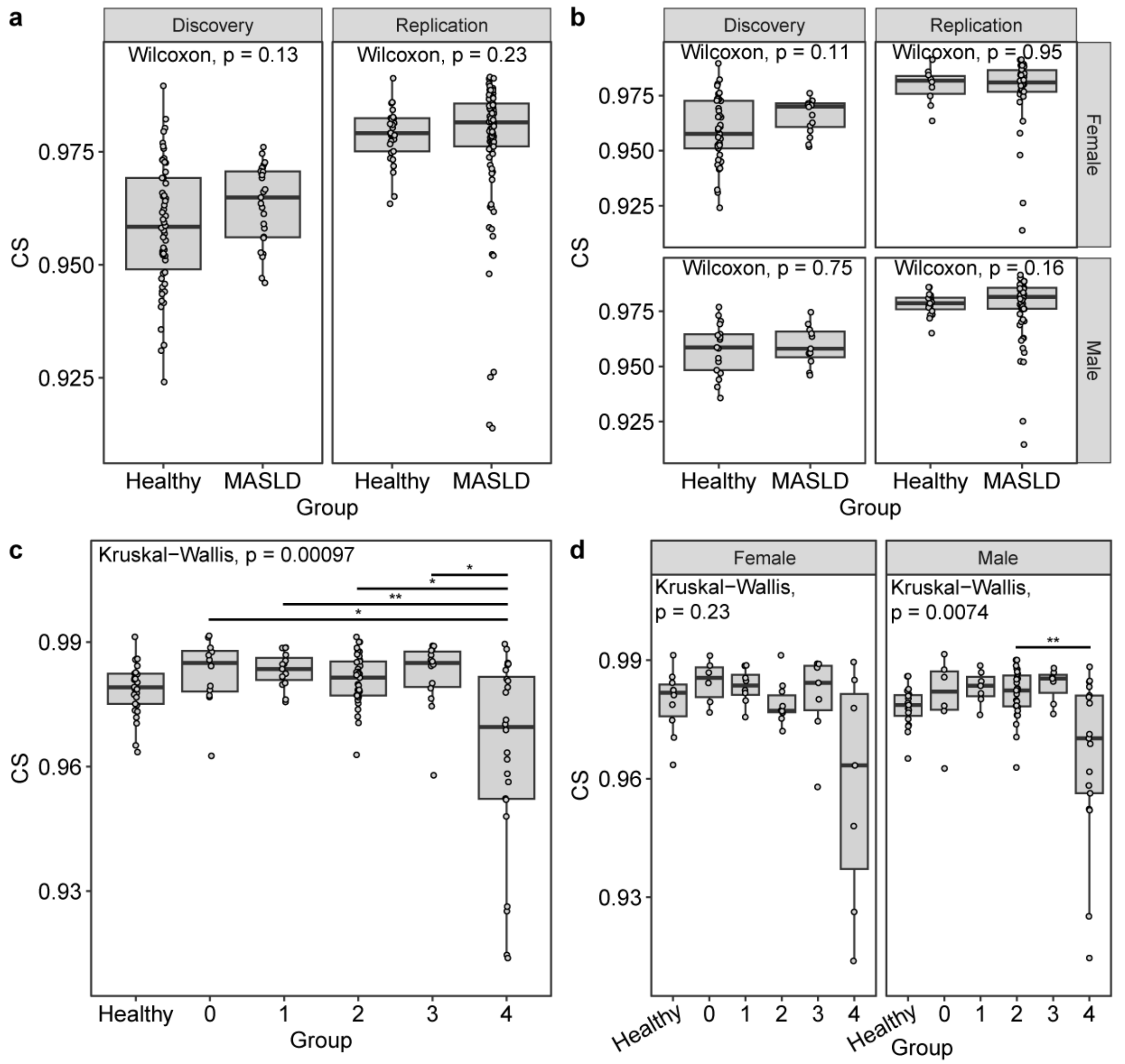

**Supplementary Figure 5. Comparison of overall sialylation (CS) between the disease groups per cohorts and sex. (a)** Differences in the relative levels of CS in healthy controls versus NAFLD patients overall, and **(b)** sex stratified in the cohorts. **(c)** Overall and **(d)** sex stratified differences in CS between healthy controls and patients with various degree of fibrosis (Brunt) in the replication cohort. While statistical testing suggested significant differences between the groups **(c-d)**, this effect was explicit for patients with advanced fibrosis (F4; cirrhosis), and no differences were observed between healthy and/or F0 patients vs. those with F1-F3 fibrosis. These results suggest that the observed fibrosis-specific effect is driven by a shift in the relative abundance of sialic acid linkages **(Supplementary Figure 2 and 6)**, rather than alterations in the relative levels of CS.

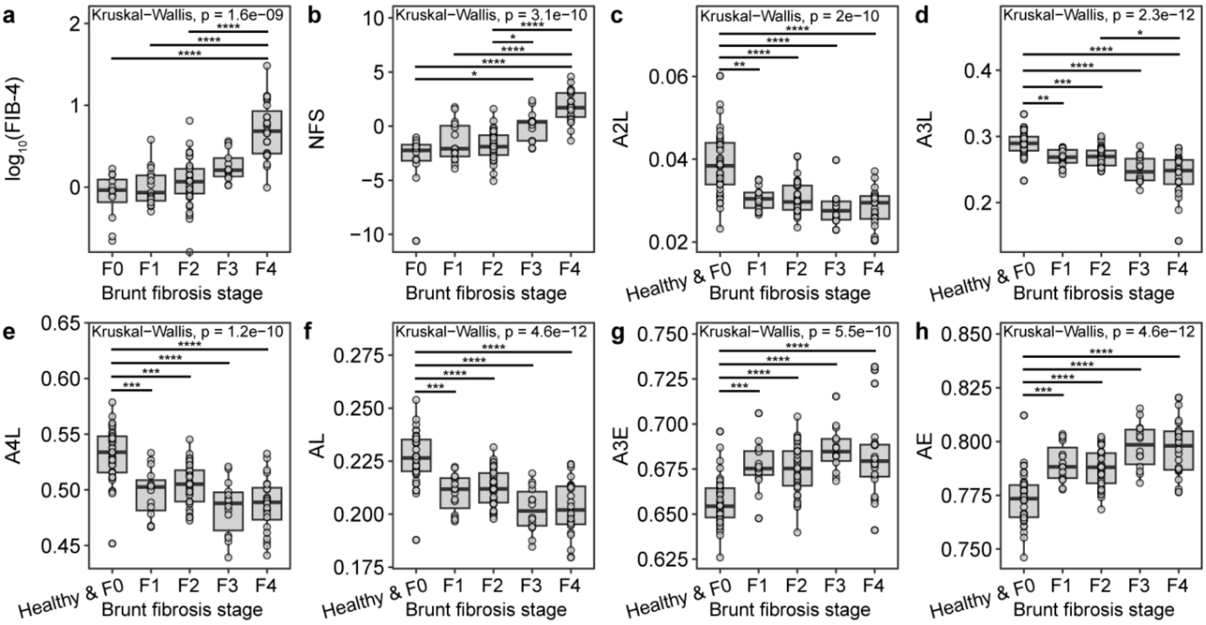

**Supplementary Figure 6. Comparison of conventional diagnostic markers and glycosylation signatures per Brunt fibrosis stage. (a-b)** Differences in the levels of FIB-4 and NFS and **(c-h)** relative levels of glycosylation signatures in F0 patients **(a-b)** pooled with healthy controls **(c-h)** vs. patients with various degree of fibrosis (Brunt). Unlike linkage-specific sialylation levels, the conventional diagnostic markers FIB-4 and NFS only indicated advanced fibrosis based on univariate statistical analysis in this cohort. Note that FIB-4 and NFS scores were not available for the healthy control group.

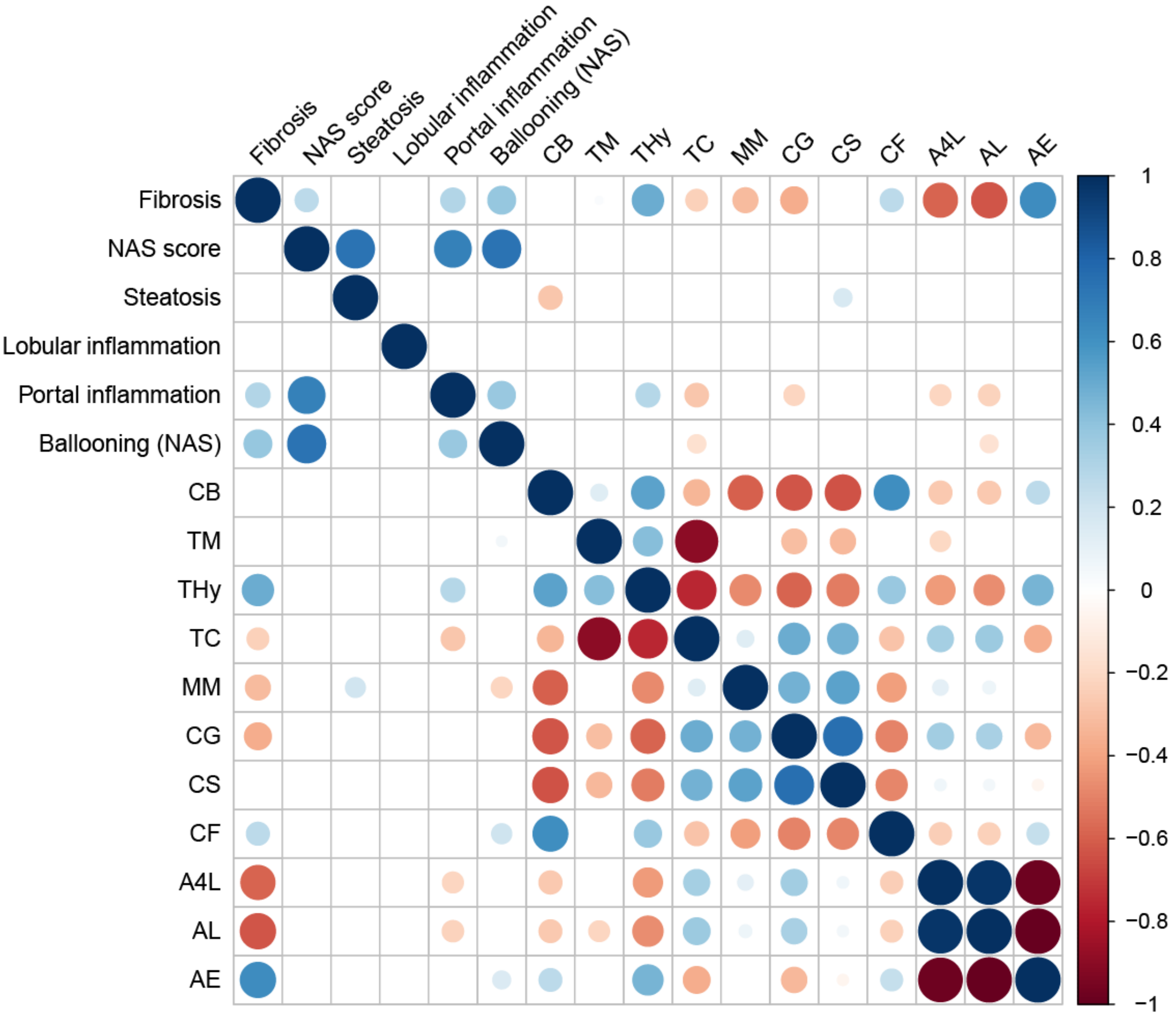

**Supplementary Figure 7. Heatmap showing significant correlations between glycosylation traits and various histological readouts including NAFLD activity score (NAS).** Color gradient: Spearman's correlation coefficient. Blank squares: insignificant correlations. Glycosylation, in particular linkage-specific sialylation (A4L, AL, AE), strongly correlates with fibrosis, but not with NAS and its individual components. Note that fibrosis additionally shows a strong positive correlation with the glycosylation trait describing the relative levels of hybrid-type glycans within all detected glycans (THy): as Brunt fibrosis score increases, so does THy. Multi-collinearity between the glycosylation traits A4L and AL/AE, is also noteworthy.
